# Transitions to Comfort Measures Only Among Patients Hospitalized With Sepsis: Clinical Drivers and Implications for Mortality

**DOI:** 10.64898/2026.10.07.26364978

**Authors:** Hannah H. Laibinis, Wei Qi, Kastin Pan, Shilei Li, Con Moran, Erin Connolly, Sarah Naz-McLean, David Regan, Grace Valentine, Ella Woehl, Elene Chamberlain, Mary Kate Cloonan, Sam Hwang, Joshua Pickard, Jacques Deguine, Deborah T. Hung, Lisa Cosimi, Ann E. Woolley

## Abstract

**Background:** Mortality in sepsis is often considered a direct consequence of infection and organ dysfunction, although many deaths occur after withdrawal of life sustaining treatment. The clinical factors leading to these decisions and their implications for interpreting sepsis mortality are not well characterized.

**Methods:** We performed a secondary analysis of a prospective observational cohort of adults with physician adjudicated sepsis at a tertiary academic medical center. Patients who transitioned to comfort measures only (CMO) during the index hospitalization underwent structured chart review to characterize baseline health and functional status, clinical severity, care preferences, anticipated outcomes following survival, and the primary reason for CMO transition.

**Results:** Among 239 patients with adjudicated sepsis, 42 (17.6%) transitioned to CMO. These patients had substantial underlying disease, including malignancy in 76% and immunocompromised status in 62%. However, 88% lived at home before hospitalization, 52% were independent in activities of daily living, an additional 26% were partially independent, and 90% were full code on admission. Progressive multi organ failure (19%), refractory septic shock (19%), and irreversible respiratory failure (12%) accounted for half of CMO transitions, while advanced frailty (5%) and poor baseline functional status (2%) were infrequent primary reasons. Median time from admission to CMO was 16 days (IQR 7 to 33), and median time from admission to death was 18 days (IQR 11 to 35). Anticipated ventilator dependence (62%), dialysis dependence (36%), and cognitive impairment frequently influenced goals of care discussions. Overall, 38 patients died during the index hospitalization, and 35 (92%) of these deaths followed transition to CMO.

**Conclusions:** Most CMO transitions in this cohort occurred after an evolving course of severe sepsis and were driven primarily by progressive organ dysfunction and failure to recover rather than advanced frailty or poor baseline function. These findings identify failure to recover after the acute septic insult as an important clinical trajectory and highlight the need to account for treatment limitation when interpreting sepsis mortality. Understanding why some patients recover from sepsis associated organ dysfunction while others progress to irreversible organ failure may help identify patients and mechanisms for future therapeutic intervention.

## Introduction

Sepsis remains one of the leading causes of hospitalization and death worldwide and is responsible for substantial morbidity, mortality, and healthcare utilization.^1,2^ Despite advances in early recognition, antimicrobial therapy, source control, and organ support, mortality among patients hospitalized with sepsis remains high, particularly among critically ill patients.^1,3^ The heterogeneity of the sepsis syndrome complicates the interpretation of mortality as an outcome and has motivated efforts to identify clinically and biologically distinct sepsis phenotypes.^4,5^

Studies of sepsis mortality have traditionally focused on physiologic severity, organ dysfunction, and comorbidity burden. Death may occur rapidly from refractory shock or fulminant organ failure. In other patients, initial stabilization is followed by persistent or progressive organ dysfunction and failure to recover. Death may then occur after a decision to withdraw life sustaining treatment. The decision to transition to comfort measures only (CMO) during hospitalization for sepsis is complex. These decisions may reflect progression of the acute illness, preexisting comorbidities, baseline functional status, anticipated quality of life following survival, and patient values and preferences. Surrogate decision makers and clinicians also contribute to these decisions through assessments of prognosis, reversibility of organ dysfunction, and the likelihood of meaningful recovery. As a result, mortality among patients with sepsis may reflect not only the severity of infection and organ dysfunction, but also decisions regarding whether continued life sustaining treatment remains appropriate and consistent with the patient’s goals.

Failure to account for transitions to comfort focused care may complicate the interpretation of sepsis mortality. Patients who transition to CMO may differ from those who continue life sustaining treatment with respect to acute illness severity, underlying disease, functional status, and goals of care. Furthermore, death after withdrawal of life sustaining therapy represents a clinically distinct pathway from death despite continued maximal treatment. Identifying patients who survive the initial septic insult but develop persistent organ dysfunction may also define a population in whom there is time to identify the mechanisms underlying failure to recover and potentially intervene. Understanding the circumstances leading to CMO may therefore provide important context for mortality as an outcome in sepsis studies.

Few studies have systematically examined why patients hospitalized with sepsis transition to comfort measures.^6^ We sought to characterize patients with sepsis who transitioned to CMO during their index hospitalization, identify the clinical and patient centered factors underlying these decisions, and determine the primary reasons for withdrawal of life sustaining therapy. We further sought to understand how transitions to CMO contribute to observed mortality in patients hospitalized with sepsis.

## Methods

### Study design and setting

The Center for Integrated Solutions to Infectious Diseases (CISID) sepsis cohort is a prospective observational study conducted at Brigham and Women’s Hospital, a tertiary academic medical center in Boston, Massachusetts. Adults with suspected sepsis were enrolled from the emergency department, inpatient units, and intensive care units between October 27, 2023, and April 15, 2026. The study was approved by the Mass General Brigham Human Research Committee institutional review board. All participants or their health care proxies provided written informed consent.

### Eligibility and enrollment

Eligible patients were adults 18 years of age or older with suspected infection, defined by a new antibiotic order together with a microbiologic culture or polymerase chain reaction test obtained within the preceding 24 hours and meeting at least two systemic inflammatory response syndrome criteria.

Potential participants were identified through a multistep screening process. Automated electronic health record reports identified potentially eligible patients across the emergency department and inpatient units. Trained study staff then performed structured chart review for evidence supporting infection or sepsis, including positive microbiologic testing, radiographic evidence of infection, or documentation of confirmed or strongly suspected infection by the primary, intensive care, or infectious diseases team. The study team subsequently discussed potential participants with the treating clinical team before approaching the patient or health care proxy for consent.

### Clinical adjudication

Enrolled participants underwent structured physician adjudication after sufficient clinical follow up to assess the hospitalization, microbiologic results, imaging findings, and response to treatment. Two physicians independently determined the presence or absence of infection and sepsis according to Sepsis-3 criteria, the presence of septic shock, and the anatomic and microbial source of infection. Discordant assessments were reviewed by a third physician. This paper’s analysis was restricted to participants with adjudicated sepsis.

### Data collection and outcomes

Demographics, comorbidities, medications, microbiologic data, and details of the index hospitalization were abstracted from the electronic health record into REDCap. Measures of acute illness included Sequential Organ Failure Assessment (SOFA), quick Sequential Organ Failure Assessment (qSOFA), and National Early Warning Score 2 (NEWS2) scores, intensive care unit (ICU) admission, duration of ICU care, shock, mechanical ventilation, vasopressor requirements, renal replacement therapy, and lactate. Clinical outcomes included hospital length of stay, in hospital mortality, 30-day mortality, 90-day mortality, and discharge disposition. Transition to CMO during the index hospitalization was identified through review of the medical record.

### Characterization of transitions to comfort measures only

For participants who transitioned to CMO, a structured retrospective chart review was performed to characterize the circumstances surrounding the decision. A predefined abstraction framework was developed to reflect factors commonly considered during clinical goals of care discussions. These included the trajectory of the acute illness, baseline health and functional status, patient preferences, anticipated treatment burden, expected quality of life following survival, complications during hospitalization, and clinician assessments of prognosis.

A single primary reason for CMO transition was assigned based on the documentation surrounding the decision and with two independent adjudicators. Prespecified categories included refractory septic shock, progressive multi organ failure, irreversible respiratory failure, catastrophic neurologic injury, advanced baseline frailty, poor baseline functional status, advanced chronic disease, patient preference or advance directive, treatment escalation inconsistent with patient goals, hospital complication, and poor predicted likelihood of recovery.

Clinical characteristics at the time of CMO transition were also abstracted, including location of care, lactate, vasopressor requirements, mechanical ventilation, renal replacement therapy, and cardiac function. Baseline residence and dependence in activities of daily living were recorded. The primary medical decision maker was categorized as the patient, health care proxy, or health care proxy making decisions based on previously expressed patient preferences. Anticipated quality of life following survival was assessed through documentation regarding expected permanent ventilator dependence, permanent dialysis dependence, and cognitive impairment.

### Statistical analysis

Cohort characteristics, infection characteristics, measures of clinical severity, and outcomes were summarized for the overall cohort and stratified by transition to CMO during the index hospitalization. Characteristics of patients who transitioned to CMO were further summarized according to baseline status, clinical status at the time of transition, decision making, anticipated quality of life following survival, and primary reason for transition. Continuous variables are reported as medians with interquartile ranges. Categorical variables are reported as counts and percentages. Analyses were descriptive. All statistical analyses were performed using R version 4.5.2

## Results

### Cohort characteristics

Among 239 enrolled patients with adjudicated sepsis, 42 patients (17.6%) transitioned to CMO during the index hospitalization (Table 1). Patients who transitioned to CMO had a median age of 67 years compared with 63 years among patients who did not transition to CMO. Malignancy was present in 76% of patients who transitioned to CMO compared with 57% of those who did not. Solid tumors were present in 57% compared with 42%, and hematologic malignancy in 26% compared with 19%. Sixty-two percent of patients who transitioned to CMO were immunocompromised compared with 47% of patients whose care was not withdrawn. A prior history of stroke was also more common in the CMO group, at 26% compared with 11%. In contrast, the prevalence of chronic renal insufficiency, pulmonary disease, autoimmune disease, and solid organ transplantation was similar between groups.

**Table 1.** Baseline characteristics of patients with adjudicated sepsis according to transition to comfort measures only. Demographic characteristics, comorbidities, immunocompromised status, and healthcare exposures before the index hospitalization are shown for the overall cohort and stratified by whether patients transitioned to CMO during the index hospitalization. Continuous variables are reported as median (IQR) and categorical variables as n (%). BMI indicates body mass index; CMO, comfort measures only; LOS, length of stay.

| Cohort Characteristics |  |  |  |
| --- | --- | --- | --- |
|  | Total<br>N = 239<br>n (%) | Care Withdrawn<br>N = 42<br>n (%) | Care Not<br>Withdrawn<br>N = 197<br>n (%) |
| **Characteristics** |  |  |  |
| Age (years) - median (IQR) | 64 (50, 74) | 67 (53, 75) | 63 (50, 73) |
| BMI (kg/m<sup>2</sup>) - median (IQR) | 28 (24, 32) | 27 (23, 31) | 28 (24, 32) |
| Sex assigned at birth |  |  |  |
| Male | 147 (62) | 32 (76) | 115 (58) |
| Female | 92 (38) | 10 (24) | 82 (42) |
| Race/Ethnicity |  |  |  |
| White Non-Hispanic | 173 (72) | 33 (79) | 140 (71) |
| Black Non-Hispanic | 23 (10) | 3 (7) | 20 (10) |
| Hispanic or Latinx | 19 (8) | 3 (7) | 16 (8) |
| Asian Non-Hispanic | 10 (4) | 1 (2) | 9 (5) |
| Other | 2 (1) | 0 (0) | 2 (1) |
| Unknown | 12 (5) | 2 (5) | 10 (5) |
| **Comorbidities** |  |  |  |
| Cardiac disease | 87 (36) | 18 (43) | 69 (35) |
| Hypertension | 155 (65) | 30 (71) | 125 (63) |
| Diabetes | 89 (37) | 17 (40) | 72 (37) |
| Stroke | 33 (14) | 11 (26) | 22 (11) |
| Pulmonary disease | 115 (48) | 18 (43) | 97 (49) |
| Autoimmune disease | 44 (18) | 7 (17) | 37 (19) |
| Cirrhosis | 17 (7) | 4 (10) | 13 (7) |
| Chronic renal insufficiency | 89 (37) | 15 (36) | 74 (38) |
| Dialysis | 16 (7) | 4 (10) | 12 (6) |
| Malignancy | 145 (61) | 32 (76) | 113 (57) |
| Solid tumor | 107 (45) | 24 (57) | 83 (42) |
| On treatment | 63 (26) | 14 (33) | 49 (25) |
| Hematologic malignancy | 48 (20) | 11 (26) | 37 (19) |
| On treatment | 39 (16) | 10 (24) | 29 (15) |
| Stem cell transplant | 19 (8) | 4 (10) | 15 (8) |
| Solid organ transplant | 18 (8) | 3 (7) | 15 (8) |
| Currently immunocompromised* | 121 (51) | 26 (62) | 95 (48) |
| Moderate to severe** | 74 (31) | 16 (38) | 58 (29) |
| **Pre-Hospitalization Exposures** |  |  |  |
| Recent surgery <30 days | 57 (24) | 11 (26) | 46 (23) |
| Prior hospitalization <30 days | 66 (28) | 16 (38) | 50 (25) |
| Infection related | 27 (11) | 6 (14) | 21 (11) |
| Sepsis related | 11 (5) | 3 (7) | 8 (4) |
| Prior hospitalization <180 days | 155 (65) | 29 (69) | 126 (64) |
| Infection related | 94 (39) | 17 (40) | 77 (39) |
| Sepsis related | 42 (18) | 6 (14) | 36 (18) |
| Total pre-admission LOS | 5 (0, 24) | 6 (0, 25) | 5 (0, 23) |
| Abbreviations: BMI = Body mass index. LOS = Length of stay. |  |  |  |
| *Based on CDC 2025 COVID-19 Vaccination Guidance for People Who Are Immunocompromised.<sup>10</sup> |  |  |  |
| **Absolute neutrophil count < 500 cells/mm<sup>3</sup> after chemotherapy; active hematologic malignancy, stem cell or solid organ transplant and on immunosuppression, prednisone equivalent > 20mg for > 2 weeks, receiving B cell inhibitor, bispecific antibody or anti-TNF inhibitor, HIV with CD4 < 200 cells/mm<sup>3</sup>, and primary immunodeficiency. |  |  |  |

**Table 2.** Anatomic and microbial sources of sepsis according to transition to comfort measures only. Anatomic sources of infection and identified microbial etiologies are shown for the overall cohort and stratified by whether patients transitioned to CMO during the index hospitalization. Microbial organisms were identified by culture or polymerase chain reaction. Categories of microbial etiology are not mutually exclusive in patients with polymicrobial infection. CMO indicates comfort measures only.

| **Infection Source** |  |  |  |
| --- | --- | --- | --- |
|  | **Total**<br>**N = 239**<br>**n (%)** | **Care Withdrawn**<br>**N = 42**<br>**n (%)** | **Care Not**<br>**Withdrawn**<br>**N = 197**<br>**n (%)** |
| **Anatomic source of sepsis** |  |  |  |
| Pulmonary | 72 (30) | 22 (52) | 50 (25) |
| Gastrointestinal | 35 (15) | 4 (10) | 31 (16) |
| Urinary | 38 (16) | 1 (2) | 37 (19) |
| Central nervous system | 3 (1) | 2 (5) | 1 (1) |
| Skin/soft tissue | 25 (10) | 4 (10) | 21 (11) |
| Hepatobiliary | 35 (15) | 6 (14) | 29 (15) |
| Vascular | 33 (14) | 7 (17) | 26 (13) |
| Bone/joint | 9 (4) | 0 (0) | 9 (5) |
| Genital | 1 (0) | 0 (0) | 1 (1) |
| **Microbial source** |  |  |  |
| Confirmed microbial source of sepsis* | 167 (70) | 28 (67) | 139 (71) |
| Gram negative | 92 (38) | 13 (31) | 79 (40) |
| Gram positive | 83 (35) | 13 (31) | 70 (36) |
| Anaerobe | 13 (5) | 1 (2) | 12 (6) |
| Fungal | 9 (4) | 5 (12) | 4 (2) |
| Viral | 13 (5) | 2 (5) | 11 (6) |
| Polymicrobial | 45 (19) | 6 (14) | 39 (20) |
| Staph aureus | 33 (14) | 7 (17) | 26 (13) |
| E. coli | 39 (16) | 2 (5) | 37 (19) |
| Enterococcus spp. | 22 (9) | 3 (7) | 19 (10) |
| Pseudomonas aeruginosa | 14 (6) | 4 (10) | 10 (5) |
| Klebsiella spp. | 28 (12) | 5 (12) | 23 (12) |
| Enterobacter spp. | 8 (3) | 1 (2) | 7 (4) |
| Candida spp. | 7 (3) | 3 (7) | 4 (2) |
| *Microbial organisms identified by culture or polymerase chain reaction (PCR). |  |  |  |

**Table 3.** Clinical severity and outcomes according to transition to comfort measures only. Clinical severity at study entry, ICU utilization, shock, mortality, and discharge disposition are shown for the overall cohort and stratified by whether patients transitioned to CMO during the index hospitalization. Continuous variables are reported as median (IQR) and categorical variables as n (%). ICU indicates intensive care unit; CMO, comfort measures only; NEWS2, National Early Warning Score 2; qSOFA, quick Sequential Organ Failure Assessment; SOFA, Sequential Organ Failure Assessment.

| **Clinical Severity and Outcomes** |  |  |  |
| --- | --- | --- | --- |
|  | **Total**<br>**N = 239**<br>**n (%)** | **Care Withdrawn**<br>**N = 42**<br>**n (%)** | **Care Not**<br>**Withdrawn**<br>**N = 197**<br>**n (%)** |
| **Status at Entry** |  |  |  |
| Entry location |  |  |  |
| Floor | 31 (13) | 4 (10) | 27 (14) |
| ICU | 166 (69) | 37 (88) | 129 (65) |
| ED | 42 (18) | 1 (2) | 41 (21) |
| Entry QSOFA score | 2 (2, 2) | 2 (2, 2) | 2 (2, 2) |
| Entry NEWS 2 score | 10 (8, 12) | 12 (10, 14) | 10 (8, 12) |
| Entry SOFA score | 9 (7, 12) | 12 (9, 15) | 9 (6, 11) |
| **Hospital Outcomes** |  |  |  |
| Length of stay (Median, IQR) | 13 (6, 28) | 16 (7, 29) | 12 (6, 27) |
| Admitted to ICU | 190 (79) | 40 (95) | 150 (76) |
| Days in ICU (Median, IQR) | 3 (1, 7) | 6 (3, 15) | 3 (1, 6) |
| Shock | 112 (47) | 26 (62) | 86 (44) |
| **Mortality** |  |  |  |
| In-hospital mortality | 38 (16) | 35 (83) | 3 (2) |
| Care withdrawn | 35 (92) | 35 (100) | 0 (0) |
| 30-day mortality | 45 (19) | 37 (88) | 8 (4) |
| 90-day mortality | 69 (29) | 42 (100) | 27 (14) |
| Care withdrawn | 61 (88) | 42 (100) | 19 (70) |

**Table 4.** Clinical characteristics and factors associated with transition to comfort measures only. Clinical status at the time of CMO transition, baseline functional status, care preferences, clinical trajectory, anticipated quality of life following survival, and the primary reason for CMO transition are shown among the 42 patients who transitioned to CMO. A single primary reason for transition was assigned through structured chart review. Continuous variables are reported as median (IQR) and categorical variables as n (%). ADL indicates activities of daily living; CMO, comfort measures only; DNR, do not resuscitate; DNI, do not intubate; HCP, health care proxy; ICU, intensive care unit; LTAC, long term acute care hospital; LV EF, left ventricular ejection fraction; QOL, quality of life; RV, right ventricular.

| **CMO Transition** |  |
| --- | --- |
|  | **Total**<br>**N = 42**<br>**n (%)** |
| **Clinical Severity** |  |
| Location Prior to Code Switch |  |
| ICU | 31 (74) |
| Floor | 11 (26) |
| Last Lactate (mmol/L) - median (IQR) | 2 (1, 3) |
| Number of Vasopressors - median (IQR) | 1 (0, 3) |
| Mechanical Ventilation | 19 (45) |
| Dialysis | 11 (26) |
| Most Recent LV EF - median (IQR) | 60 (55, 65) |
| Moderate to Severe RV Dysfunction | 4 (10) |
| Severe Valvular Disease | 2 (5) |
| **Baseline Status** |  |
| Age (years) - median (IQR) | 67 (53, 75) |
| Residence before Hospitalization |  |
| Home | 37 (88) |
| Rehab/LTAC | 5 (12) |
| ADL Dependence |  |
| Independent | 22 (52) |
| Partially Independent | 11 (26) |
| Not Independent | 9 (21) |
| **Care Preferences** |  |
| Full Code on Admission | 38 (90) |
| Primary Medical Decision Maker |  |
| Patient | 6 (14) |
| HCP | 20 (48) |
| HCP w/ prior pt preferences | 16 (38) |
| **Care Trajectory** |  |
| Admission to Care Withdrawn - median days (IQR) | 16 (7, 33) |
| Admission to Death - median days (IQR) | 18 (11, 35) |
| Sepsis 'Entry' to Care Withdrawn - median days (IQR) | 7 (4, 18) |
| Sepsis 'Entry' to Death - median days (IQR) | 12 (5, 20) |
| **Perceived QOL After Survival** |  |
| Permanent Dialysis | 15 (36) |
| Ventilator Dependence | 26 (62) |
| Cognitive Impairment |  |
| Mild | 6 (14) |
| Severe | 7 (17) |
| **Primary CMO Reason** |  |
| Advanced baseline frailty | 2 (5) |
| Advanced chronic disease | 7 (17) |
| Catastrophic neurologic injury | 1 (2) |
| Hospital complication | 1 (2) |
| Irreversible respiratory failure | 5 (12) |
| Patient preference / advance directive | 4 (10) |
| Poor baseline functional status | 1 (2) |
| Poor predicted likelihood of recovery | 1 (2) |
| Progressive multi-organ failure | 8 (19) |
| Refractory septic shock | 8 (19) |
| Treatment escalation inconsistent with goals | 4 (10) |

Recent healthcare exposure was common in both groups. Thirty eight percent of patients who transitioned to CMO had been hospitalized during the preceding 30 days compared with 25% of those who did not. Hospitalization within the preceding 180 days occurred in 69% and 64%, respectively.

Pulmonary infection was more common among patients who transitioned to CMO, accounting for 52% of infections compared with 25% among patients who did not. Urinary infection was less common, occurring in 2% compared with 19%. A microbial etiology was identified in 67% of the CMO group and 71% of patients who did not transition to CMO.

### Severity of illness and outcomes

Patients who ultimately transitioned to CMO had greater acute illness severity at study entry. Median SOFA score was 12 compared with nine among patients who did not transition to CMO. Median NEWS2 score was 12 compared with 10. Ninety five percent of patients who transitioned to CMO were admitted to the ICU compared with 76% of those who did not. Shock occurred in 62% compared with 44%. Patients who transitioned to CMO spent a median of six days in the ICU compared with three days among those who did not. Median hospital length of stay was 16 days compared with 12 days.

Overall, 38 of 239 patients (16%) died during the index hospitalization. Thirty five of these 38 deaths (92%) occurred among patients whose care was withdrawn. In hospital mortality was 83% among patients who transitioned to CMO compared with 2% among those who did not.

### Clinical status at transition to CMO

Among the 42 patients who transitioned to CMO, 31 (74%) were in the ICU at the time of transition and 11 (26%) were on an inpatient floor. The median lactate immediately preceding transition was 2 mmol/L (IQR 1 to 3), and patients were receiving a median of one vasopressor (IQR 0 to 3). Nineteen patients (45%) were receiving mechanical ventilation and 11 (26%) were receiving renal replacement therapy. Despite the severity of illness, cardiac dysfunction was uncommon. The median left ventricular ejection fraction was 60% (IQR 55, 65), only four patients (10%) had moderate or severe right ventricular dysfunction, and severe valvular disease was present in two patients (5%).

The median time from hospital admission to transition to CMO was 16 days (IQR 7, 33). The median time from sepsis study entry to transition to CMO was seven days (IQR 4, 18). Median time from admission to death was 18 days (IQR 11, 35), and median time from sepsis study entry to death was 12 days (IQR 5, 20).

### Baseline health and functional status

Although patients who transitioned to CMO had substantial comorbidity, most had relatively preserved baseline functional status. Thirty-seven patients (88%) lived at home before hospitalization. Twenty-two (52%) were independent in activities of daily living and an additional 11 (26%) were partially independent. Only nine patients (21%) were not independent. Thirty-eight patients (90%) were full code at hospital admission, while only four (10%) had a preexisting DNR or DNI order.

### Decision making and anticipated quality of life following survival

Only six patients (14%) were the primary decision makers for the transition to CMO. A health care proxy was the primary decision maker in 20 cases (48%). In an additional 16 cases (38%), the health care proxy made decisions based on previously expressed patient preferences.

Concerns regarding anticipated quality of life following survival were common. Permanent ventilator dependence was anticipated in 26 patients (62%), and permanent dialysis dependence in 15 (36%). Severe cognitive impairment was anticipated in seven patients (17%), with an additional six (14%) expected to have cognitive impairment that was not considered severe.

### Primary reasons for transition to CMO

Acute irreversible organ failure was the most frequent driver of transition to CMO. Progressive multi organ failure and refractory septic shock were each identified as the primary reason in eight patients (19%). Irreversible respiratory failure was the primary reason in five patients (12%). Together, these acute physiologic causes accounted for 21 of 42 transitions (50%).

Advanced chronic disease was the primary reason for CMO transition in seven patients (17%). Patient preference or a prior advance directive accounted for four transitions (10%), and treatment escalation considered inconsistent with patient goals accounted for an additional four (10%). Advanced baseline frailty was the primary reason in only two patients (5%), while poor baseline functional status was the primary reason in one patient (2%). Catastrophic neurologic injury, hospital complication, and poor predicted likelihood of recovery each accounted for one transition.

## Discussion

In this prospective cohort of patients with adjudicated sepsis, nearly one in five transitioned to comfort focused care during the index hospitalization, accounting for 92% of in-hospital deaths. CMO most often followed failure to recover from severe acute illness. Progressive multi organ failure, refractory septic shock, and irreversible respiratory failure accounted for half of all transitions. In contrast, advanced frailty and poor baseline functional status were rarely the primary reasons for CMO.

Although patients who transitioned to CMO had substantial medical comorbidity, most had relatively preserved baseline function. Malignancy was present in 76% of patients who transitioned to CMO compared with 57% of those who did not, and 62% were immunocompromised compared with 47%. Despite this burden of underlying disease, 88% lived at home, 52% were independent in activities of daily living, and another 26% were partially independent. Ninety percent were full code on admission. Severe cardiac dysfunction was also uncommon. These findings suggest that underlying disease contributed to prognosis, but CMO was generally not driven by poor baseline function or advanced chronic organ failure alone. Rather, patients developed severe sepsis associated organ dysfunction from which they failed to recover.

The timing of CMO further supports this trajectory. The median time from admission to CMO was 16 days and the median time from admission to death was 18 days. These patients generally survived the initial phase of sepsis and received active treatment before the decision to transition to CMO. At that point, anticipated outcomes following survival were also important. Nearly two thirds were expected to remain ventilator dependent, more than one third were expected to require permanent dialysis, and cognitive impairment was a concern in approximately one third. Thus, decisions to withdraw life sustaining treatment reflected both the likelihood of recovery and whether the anticipated outcome was consistent with the patient’s goals and values.

These findings are also important for interpreting mortality in sepsis studies. Mortality is commonly treated as a single outcome, but the pathways leading to death may differ substantially. In our cohort, nearly all in hospital deaths occurred after withdrawal of life sustaining treatment. Death therefore often followed a period of persistent organ dysfunction and a subsequent decision that further treatment was unlikely to result in an acceptable recovery. Sepsis studies using mortality as an endpoint should consider capturing CMO and other limitations of life sustaining treatment to better distinguish these clinical trajectories.

Our findings also identify a potentially important population for future therapeutic studies. Patients who survive the initial septic insult but develop persistent or progressive organ dysfunction may represent a distinct group in whom there is time to identify mechanisms associated with failure to recover and potentially intervene. Sepsis is increasingly recognized as a heterogeneous syndrome, and recent studies have identified clinical and biological phenotypes associated with different outcomes and treatment responses.^4,5,7–9^ The key biological question raised by our findings is why some patients recover from sepsis associated organ dysfunction while others enter an irreversible trajectory.

Answering this question may require moving beyond prediction of mortality at presentation. Serial clinical and biological assessment during the first several days of sepsis could identify patients who are failing to recover and define the mechanisms underlying persistent organ dysfunction. This approach could inform therapies directed at promoting organ recovery and could help identify patients most likely to benefit from those therapies. Future trials could enroll patients based on an evolving clinical or biological phenotype rather than sepsis alone and test interventions before organ dysfunction becomes irreversible.

Importantly, our cohort does not adequately capture all trajectories of death from sepsis. Because enrollment required time to identify patients, obtain consent, and complete initial study blood samples, patients who died or transitioned to CMO shortly after presentation were less likely to be enrolled. Our findings primarily characterize patients who survived the earliest phase of sepsis but subsequently failed to recover. The biology of rapid early death may be different and will require studies specifically designed to capture patients during the earliest illness period.

This study has additional limitations. It was conducted at a single tertiary academic medical center and included a relatively small number of patients who transitioned to CMO. The cohort had a high prevalence of malignancy and immunocompromising conditions, which may limit generalizability. Classification of the primary reason for CMO was retrospective and required assignment of a single primary reason to decisions that were often multifactorial. Patient and surrogate perspectives were ascertained from clinical documentation rather than direct interviews.

In conclusion, transition to CMO was common among patients hospitalized with sepsis and preceded the vast majority of in-hospital deaths. Most transitions followed progressive organ dysfunction and failure to recover rather than advanced frailty or poor baseline functional status. These findings highlight failure to recover as an important clinical trajectory in sepsis and raise a central biological question: why do some patients recover from sepsis associated organ dysfunction while others progress to irreversible organ failure? Defining the mechanisms underlying this trajectory may help identify patients who remain amenable to intervention and inform the development and testing of therapies aimed at promoting recovery.

## Data Availability

All data produced in the present study are available upon reasonable request to the authors.

